# How unequal vaccine distribution promotes the evolution of vaccine escape

**DOI:** 10.1101/2021.03.27.21254453

**Authors:** Philip J Gerrish, Fernando Saldaña, Benjamin Galeota-Sprung, Alexandre Colato, Erika E Rodriguez, Jorge X Velasco Hernández

## Abstract

Health officials warn that SARS-CoV-2 vaccines must be uniformly distributed within and among countries if we are to quell the ongoing pandemic. Yet there has been little critical assessment of the underlying reasons for this warning. Here, we explicitly show why vaccine equity is necessary. Perhaps counter-intuitively, we find that vaccine escape mutants are less likely to come from highly vaccinated regions where there is strong selection pressure favoring vaccine escape and more likely to come from neighboring unvaccinated regions where there is no selection favoring escape. Unvaccinated geographic regions thus provide evolutionary reservoirs from which new strains can arise and cause new epidemics within neighboring vaccinated regions and beyond. Our findings have timely implications for vaccine rollout strategies and public health policy.

## Main text

In the face of the ongoing SARS-CoV-2 pandemic, researchers and companies around the world have vacillated between competition and cooperation in the race for a vaccine that could restore some societal and economic normalcy. As of March 2021, there are 82 vaccines in clinical development and another 182 vaccines in pre-clinical stages. These very hopeful advances happened in record time: the vaccine from the University of Oxford and the AstraZeneca vaccine took approximately 10 months to be ready for use.

Vaccine rollout and distribution are still just beginning at the time of writing this paper. In the western hemisphere, the three largest countries – the US, Brazil, and Mexico – have vaccinated 12%, 2%, and 1% of their populations, respectively [1]. Significantly, the distribution of the vaccine to date has been highly non-uniform among and within these three countries and around the globe.

As vaccines are being distributed, the emergence of vaccine escape presents a major concern [2–9]. Vaccine escape is the appearance and spread of viral variants that can infect and cause illness in vaccinated hosts. These variants can achieve this because they have acquired mutations allowing them to escape detection by antibodies created in response to vaccination. Researchers around the globe continue to identify emerging variants of SARS-CoV-2 that are increasingly refractory to vaccine-induced antibodies. These strains carry mutations in the receptor-binding domain (RBD) and the N-terminal domain (NTD) of the spike surface glycoprotein targeted by vaccines [5, 6, 8]. Put differently, vaccine escape in the ongoing pandemic is, to some degree, already occurring.

The threat of vaccine escape in the past has depended largely on the particular virus. Polio and measles vaccines are two stellar examples of highly effective vaccines to which the respective viruses have not evolved escape strains. Flu vaccines, on the other hand, are notoriously “leaky”, with rampant vaccine escape emerging every flu season and creating the need for a new flu vaccine every year. Vaccine escape has effectively prevented the development of an HIV vaccine because mutants able to “escape” any conceivable vaccine target preexist in circulating virus. Where the many different SARS-CoV-2 vaccines stand in this wide spectrum of vaccine-escape susceptibility is still a matter of debate, but increasingly the evidence indicates escape is a real threat [2–9]. The E484K mutation in the backgrounds of UK variant B.1.1.7 or South African variant B.1.135 are two particularly worrisome variants [5, 9]. There is even some concern, and evidence, that new variants may be able to evade natural immunity to SARS-CoV-2 in previously-infected hosts through “immune escape” [8–10]; this does not bode well for prospects of lasting vaccine-induced immunity [11, 12]. Finally, a recent study [13] reveals that closely-related endemic human coronavirus 229E displays evidence of “antigenic drift” – the same process of rapid antigenic evolution that occurs in Influenza.

Vaccine escape can be viewed as analogous to drug resistance. In the evolution of drug resistance, it is well-established that “privileged sites” in an infected host in which the administered drug is somehow restricted due to physiological constraints (e.g, the blood-brain barrier [14]), can play a very key role [14–16]. In such sites, the population size of the infectious agent can remain large because there is essentially no drug to suppress it. In these large sub-populations, mutants that are resistant to the drug can increase in frequency without selective constraints for or against. When resistant mutants from such privileged sites migrate back into sites with unrestricted drug concentrations, these “unprivileged sites” quickly succumb to fixation of resistant mutants. The same phenomenon is well-documented in biofilms [16–19], wherein regions of a biofilm that are shielded from antibiotics provide reservoirs in which resistance mutations evolve neutrally and can subsequently migrate to unshielded regions, rendering the antibiotic ineffective in these regions and eventually in the entire biofilm. The same principle even applies to cancer: disparities in drug concentration can promote the evolution of drug resistance [20].

The lessons learned from drug resistance point to two key factors that could facilitate the evolution of vaccine escape, namely, population size and population structure [21, 22]. Population size is important because the overwhelming majority of mutations occur during replication. Smaller populations mean fewer replications which means reduced opportunity for mutations to arise; this simple principle is the basis for health officials’ repeated pleas for continued social distancing and facemask usage (in the context of the current SARS-CoV-2 pandemic) despite the existence of vaccines. Population structure is important because local epidemics can vary in size and vaccine coverage and thus harbor the vaccine equivalent of privileged sites, mentioned above, albeit at the host-population and not within-host level. A recent study [23] looks at the effects of different kinds of population compartmentalization on the risk of vaccine escape, namely, age and vulnerability.

A further lesson from drug resistance studies derives from the observation that variability in drug distribution can have more of an impact on the evolution of resistance than overall drug concentration [21, 22]. Extrapolating to vaccine escape, this would indicate that geographic variability in vaccine distribution can pose a bigger threat of vaccine escape than factors such as public distrust and fake news that reduce vaccine participation throughout the population.

To assess claims that vaccine equity is essential, and to validate our verbal extrapolations from drug resistance evolution, we employ mathematical models and simulations of vaccine escape evolution. In our basic model, there are just two local epidemics in geographically neighboring regions or “patches”. First, we assume that one patch has access to a vaccine, the other does not, and we study how the unvaccinated patch affects the probability of vaccine escape in the vaccinated patch (Fig 1a). Second, we assume there is a limited supply of vaccine, and we study how asymmetric vs symmetric distribution to the two patches influences the probability of vaccine escape (Fig 1b).

**Fig 1.**
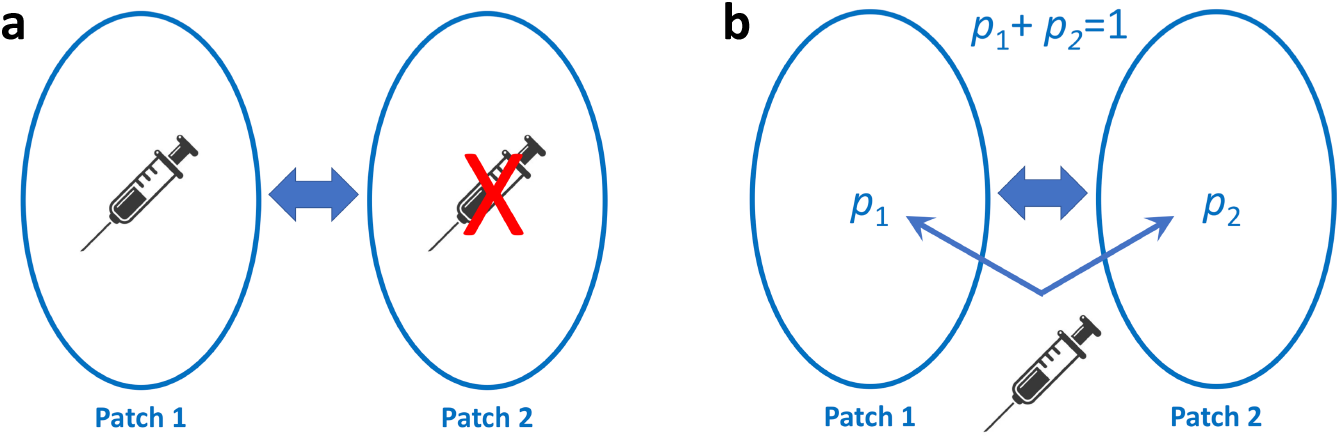
Schematic of two-patch scenarios addressed. With our two-patch model, we address two kinds of questions: **a**) How do unvaccinated regions affect vaccine efficacy in neighboring vaccinated regions? Here, we assume only Patch 1 receives vaccination. **b**) How does equal vs unequal vaccine distribution affect overall vaccine efficacy? Here, some fraction *p*_2_ of vaccine goes to Patch 2, and fraction *p*_1_ = 1 − *p*_2_ goes to Patch 1. *p*_2_ is the quantity represented by the horizontal axes in Figs 3 and 4. Maximum disparity is achieved at *p*_2_ = 0 and *p*_2_ = 1. Maximum equality is achieved at *p*_1_ = 0.5. The rate of inter-patch infection is controlled by parameter *λ* which can be interpreted as the porosity of a border separating the two regions.

We can assume that an escape mutant will always have a selective advantage in a vaccinated population (SI), simply because there is a larger number hosts it can infect (susceptible and vaccinated hosts) than the wildtype (infects susceptible hosts only). Thus, we do not need to explicitly model the transmission of and dynamics of escape mutants after they have emerged; we can focus simply on the timing of emergence of the first escape mutant. To this end, we simply model the accumulation of escape mutations from wildtype and focus on the timing of the first infection event in which a new host is infected with an escape mutant, which we will call an “escape-infection” event.

In vaccinated Patch 1, there will be strong selection for vaccine escape but limited opportunity for escape mutations to arise simply because of the reduced number of unvaccinated susceptible hosts. In unvaccinated Patch 2, escape mutations have no selective advantage, but there is a larger number of unvaccinated susceptible hosts.

Our model is described by the following equations:

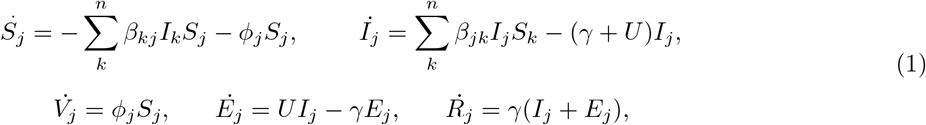

where *S*_*j*_, *V*_*j*_, *I*_*j*_, *E*_*j*_, and *R*_*j*_ are the fraction of the population that are susceptible, vaccinated, infected, infected with escape mutant, and recovered, respectively, in Patch *j*; *β*_*ij*_ is the transmission rate from Patch *i* to Patch *j* (*β*_*jj*_ is the transmission rate within Patch *j*); *ϕ*_*j*_ is vaccination rate in Patch *j*; *γ* is recovery rate; *U* is a composite per-host mutation rate from wildtype virus to escape mutant virus (see discussion below); *n* is number of patches; dots indicate time derivatives.

We note the absence of a contagion term in the equation for *E*_*j*_. This term is not needed for our purposes because our focus is only on the first escape-infection event – a discrete event. Furthermore, this term can lead to erroneous results because ours is a continuous model: a contagion term would allow for transmission to fractions of individual hosts that can erroneously amplify the vaccine escape mutant prior to the first escape-infection event.

Here, we assume there are only two patches, *n* = 2 and *j* ∈ [1, 2]. Our more complex models and detailed simulations are described in the Supplementary Information (SI).

We define random variable *T*_*ij*_ as the time of the first infection event in which a new host in Patch *j* is infected by an escape mutant that arose in Patch *i*. Such infection events occur with rate *r*_*ij*_(*t*) = *β*_*ij*_*E*_*i*_(*t*)(*S*_*j*_(*t*) + *σV*_*j*_(*t*)), where *σ* allows for varying levels of escape reflecting the observed spectrum of partial immunity against different variants ranging from no escape *σ* = 0 to full escape *σ* = 1.

For now, we will assume intra-patch transmission rates are equal, *β*_*jj*_ = *β*, and inter-patch transmission rates are equal, *β*_*ij*_|_*i*≠*j*_ = *β*_×_. We let *β*_×_ = *λβ* and we assume *λ* ≪ 1 to reflect the fact that inter-patch transmission will typically be much less frequent than intra-patch transmission. We define random variable *T*_*f*_ as the time at which the last infected individual recovers.

The three quantities of interest are: 1) *p* = 𝕡{*T*_11_ > *T*_21_ | *T*_12_ < *T*_*f*_ ∨ *T*_11_ < *T*_*f*_}, the probability that vaccine escape in Patch 1 comes not from Patch 1 but from neighboring unvaccinated Patch 2, conditioned on vaccine escape emerging in Patch 1 from one of the two patches (Fig 1a), 2) *f* = 𝕡{*T*_21_ < *T*_*f*_ ∨ *T*_11_ < *T*_*f*_}/𝕡{*T*_11_ < *T*_*f*_}, the factor by which the probability of vaccine escape in Patch 1 is increased by having neighboring unvaccinated Patch 2 (Fig 1a), and 3) *ε* = 𝕡{*T*_11_ < *T*_*f*_ ∨ *T*_12_ < *T*_*f*_ ∨ *T*_21_ < *T*_*f*_ ∨ *T*_22_ < *T*_*f*_}, the total probability of vaccine escape in the two patches as a function of vaccine distribution between the two patches (Fig 1b). These quantities are rather immediate functions of the *r*_*ij*_(*t*); they are derived in the SI and plotted in Figs 2, 3 and 4.

**Fig 2.**
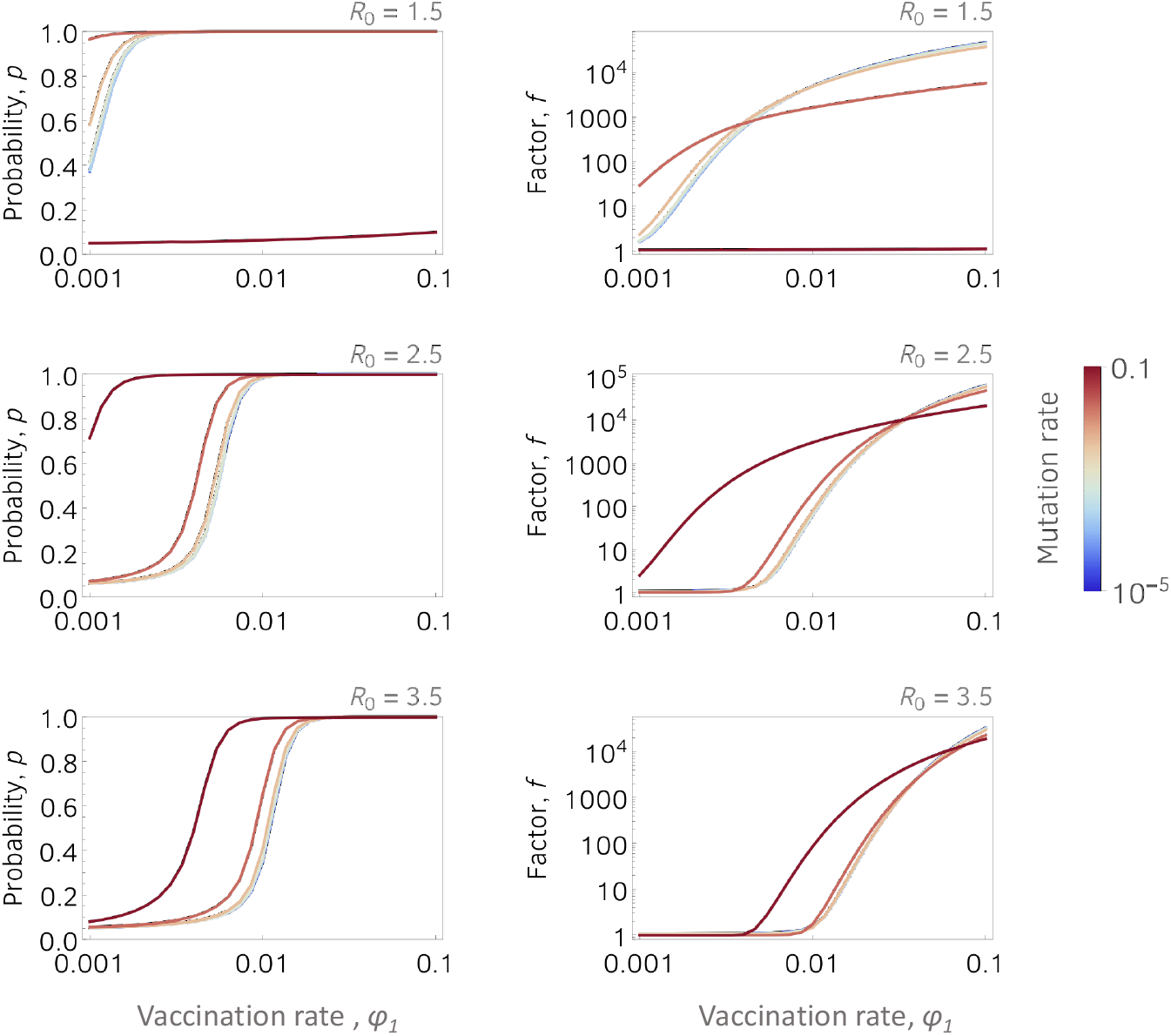
Effect of unvaccinated neighboring regions on probability of vaccine escape in vaccinated regions. We plot: *p*, the probability that vaccine escape emerging in vaccinated Patch 1 comes not from Patch 1 but from Patch 2 (*left column*), and *f*, the factor by which the probability of vaccine escape emerging in Patch 1 is increased as a consequence of having unvaccinated neighboring Patch 2 (*right column*). Horizontal axes indicate Patch 1 vaccination rate, *ϕ*_1_, and *ϕ*_2_ = 0. Parameters not specified in the plots are: *λ* = 0.05, *γ* = 0.1, *β* = *γR*_0_, and *N* = 10^5^. Colors indicate the value of the composite mutation parameter *U* as indicated by the legend. Expressions for *p* and *f* are specified in the main text and derived in the SI. For these plots, we have assumed that vaccination begins at the time the epidemic begins. Departures from this assumption as well as other explorations of parameter space are in the SI.

**Fig 3.**
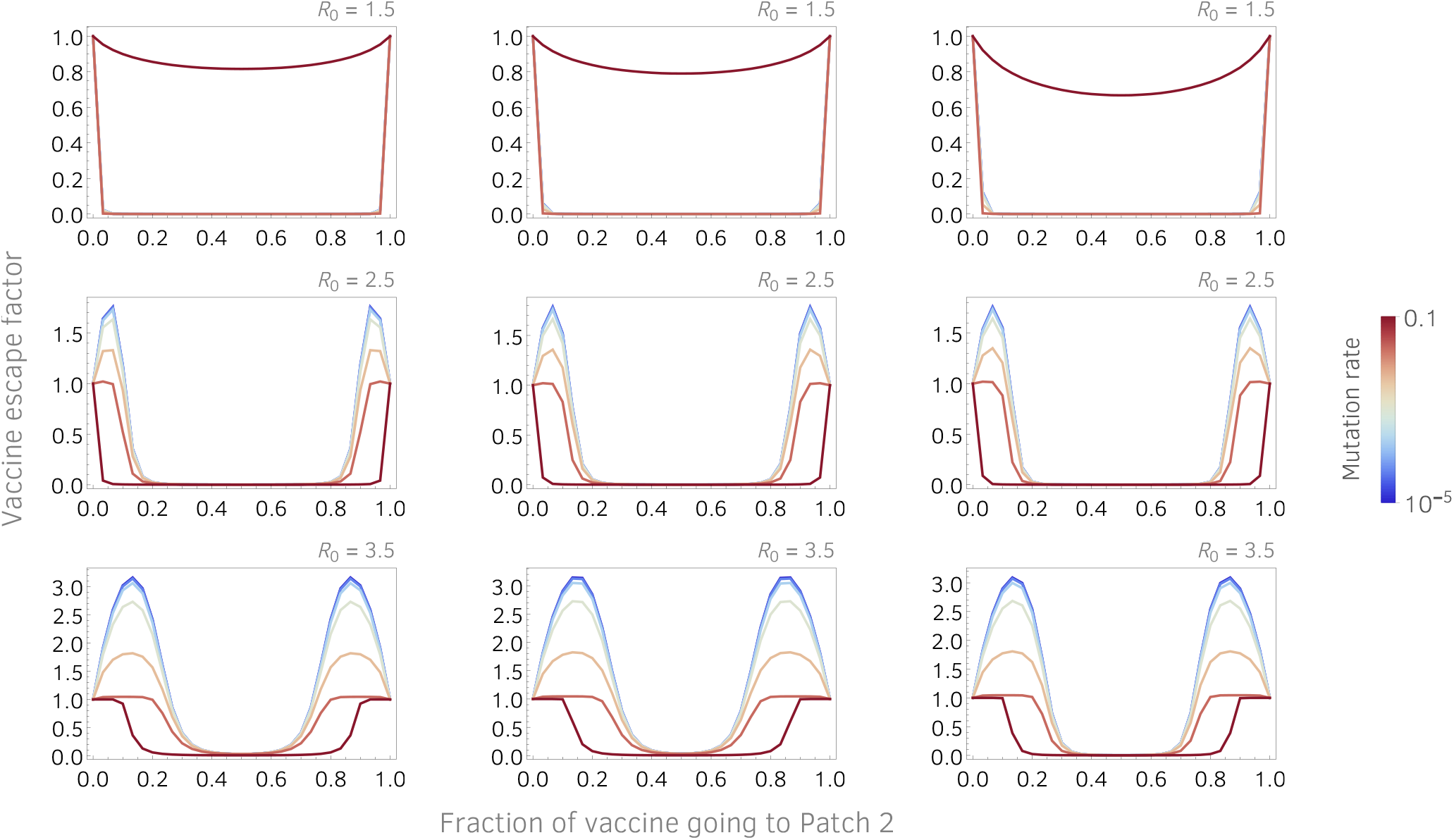
Effect of vaccine disparity on probability of vaccine escape. Vaccine escape factor is plotted as a function the fraction of vaccine that goes to Patch 2. Vaccine escape factor is defined as vaccine escape probability, *ε*, divided by the escape probability when all vaccine goes to one of the two patches (maximum disparity). Parameters are: *λ* = 0.05, *γ* = 0.1, *β* = *γR*_0_, *N* = 10^5^ and *left column*: *V* (0) = 0, *ϕ*_1_ + *ϕ*_2_ = 0.05; *middle column*: *V* (0) = 0.2, *ϕ*_1_ + *ϕ*_2_ = 0.02; *right column*: *V* (0) = 0.6, *ϕ*_1_ + *ϕ*_2_ = 0.1. Colors indicate the value of the composite mutation parameter *U* as indicated by the legend.

**Fig 4.**
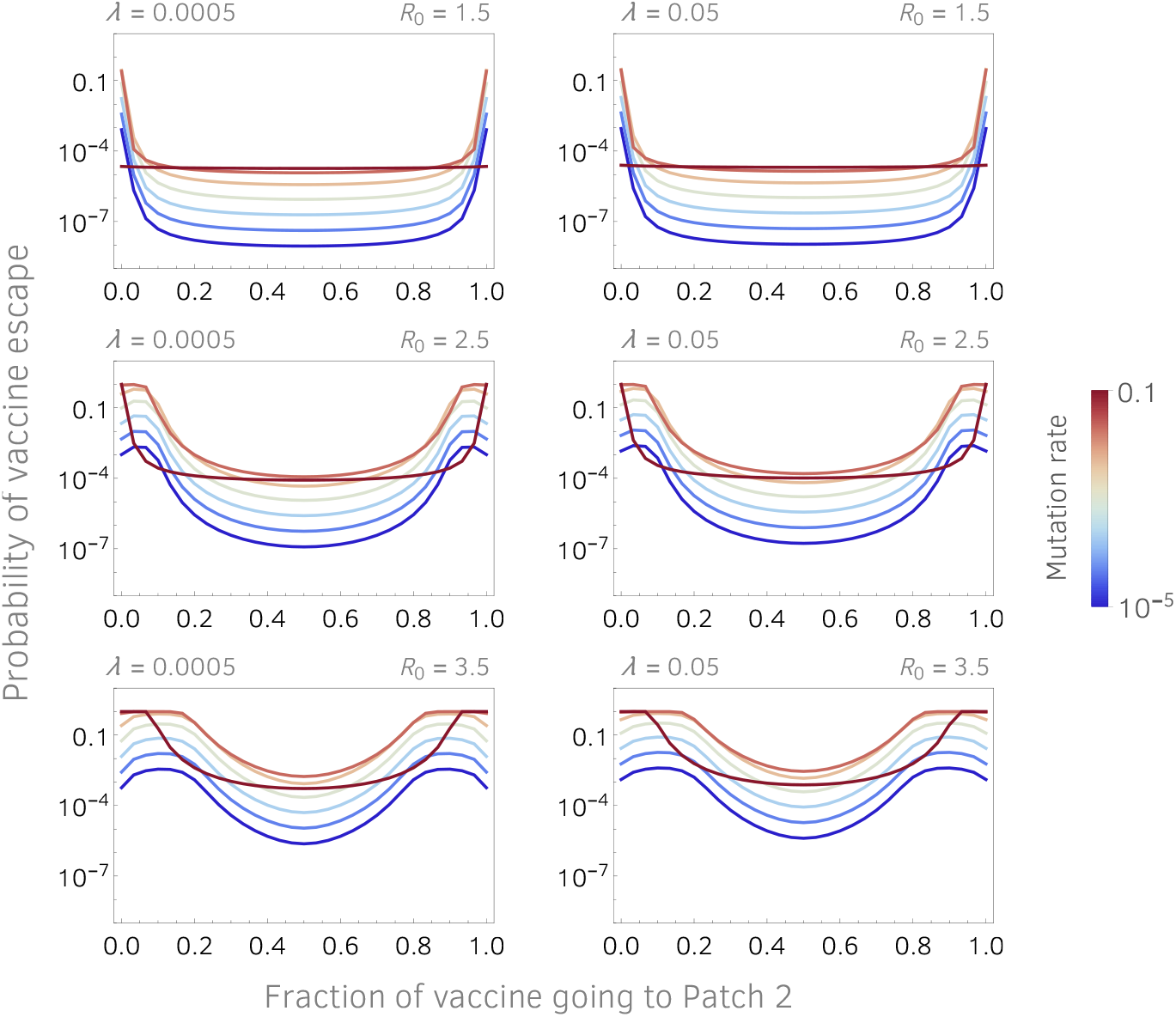
Benefit of tightening borders is small; benefit of vaccine equity is large. Probability of vaccine escape, *ε*, is plotted as a function the fraction of vaccine that goes to Patch 2. The left- and right-hand columns plot escape probabilities when border porosity is very low (*λ* = 0.0005) and much higher (*λ* = 0.05), respectively. Other parameters are: *γ* = 0.1, *β* = *γR*_0_, *N* = 10^5^ and *V* (0) = 0, *ϕ*_1_ + *ϕ*_2_ = 0.05. Colors indicate the value of the composite mutation parameter *U* as indicated by the legend.

A striking feature of Fig 2 is how large the effect of an unvaccinated neighboring region can be: the probability of vaccine escape can be orders of magnitude higher with an unvaccinated neighboring region than without it. At the time of writing this paper, an area of the world that approximates our model in a rather extreme corner of parameter space – in the context of the ongoing SARS-CoV-2 pandemic – is Israel and its neighbors: Israel presently has the highest vaccine coverage in the world [24] while its neighbors have among the lowest. Our findings would recommend vigilance for vaccine escape in this and many other areas of the world that have significant disparity in vaccine distribution both between and within countries. Figure 3 explicitly shows the effect of vaccine disparity between the two patches. Equal vaccination between the two patches gives the lowest probability of vaccine escape. Curiously, for medium to high reproductive numbers and low mutation rates, moderate disparities in vaccination can promote the emergence of vaccine escape more strongly than extreme (all or nothing) disparities.

Our parameter *U* is a composite parameter: it is the rate at which the transmission chain among hosts finally leads to one host infecting another host with a vaccine escape mutant, which we refer to above as an escape-infection event. As such, this parameter incorporates the mutation rate of the virus as well as any effect on within-host fitness it may have: a decrease (increase) in within-host fitness will effectively decrease (increase) *U*. Within-host fitness of SARS-CoV-2 should not be affected by humoral immunity of the host because transmissibility of SARS-CoV-2 peaks around the time of onset of symptoms [25], whereas a robust antibody response is not mounted until roughly ten days after the onset of symptoms [25, 26]. Thus any effect that escape mutations have on within-host fitness will not be antigenic in nature and will thus expose any pleiotropic fitness effects of vaccine escape. In this light, *U* may be viewed as primarily encapsulating two factors: viral mutation rate, and any pleiotropic fitness effect vaccine escape may have.

The parameters of our model most readily affected by public policy are *λ, β* and *ϕ*_*j*_: *λ* can be reduced, for example by reducing border porosity between regions or countries; *β* can be reduced by facemasks and social distancing, and *ϕ*_*j*_ can be made more uniform by equitable vaccine distribution. Figure 4 plots vaccine escape probabilities at values of *λ* that differ by two orders of magnitude; it is striking how little difference this makes. The implication is that reducing border porosity by a substantial amount (likely to be very costly) has a negligible effect on the risk of vaccine escape; by contrast, equal vaccine distribution (likely to be much less costly) reduces the risk of vaccine escape enormously. As vaccines become increasingly available, restrictions on movement and contact are slowly being lifted in parallel, increasing both *λ* and *β*. The relaxation of these restrictions may have adverse effects if done too quickly [27–29], but our findings would indicate that the risk of vaccine escape may not be affected much as long as the distribution of vaccine continues to be equitable.

Our findings may be understood intuitively by recalling the two ingredients required for Darwinian evolution, namely, heritable variation and natural selection. Geographic regions with low vaccine coverage are evolutionary reservoirs that provide heritable variation in the virus’s susceptibility to vaccine-induced antibodies. Geographic regions with high vaccine coverage provide the natural selection component. Regions of low and high vaccine coverage thus act in concert to promote the evolution of vaccine escape mutants that can give rise to renewed and unfettered spread of SARS-CoV-2. Our findings (SI) also show that highly granular vaccine disparities – large adjacent populations such as cities, states or countries that differ in vaccine accessibility – most effectively promote the evolution of vaccine escape. It may be that vaccine escape is inevitable and that SARS-CoV-2 will eventually become endemic [30, 31], in which case vaccine updating and exploration of new antigenic targets [32] will become the norm. Or we may have a window of opportunity now to prevent that outcome, in which case present vaccine rollout strategies could make the difference. Vaccine updating, optimizing deployment of the many different available vaccines [33] and, as we have shown, vaccine equity, are key ingredients of these strategies.

## Data Availability

Computer code for simulations is freely available upon request to the corresponding author.

## Acknowledgements

The authors thank M. Fricke, the Center for Advanced Research Computing and the Biology Department at the University of New Mexico for critical infrastructure and computational support; J. Gerrish, and D. Chencha for helpful comments.

## Author contributions

Conceptualization: PJG

Methodology: PJG, JXVH, FS, BGS, AC, ER

Investigation: PJG, JXVH, FS, BGS, AC, ER

Writing – original draft: PJG, BGS

## Data and materials availability

Computer code for simulations freely available upon request to PJG. All other methodological details are available in the main text or the supplementary information.

## Supplementary Information

Supplementary Text

Figs. S1 to S21

