## Supplemental Methods for "How unequal vaccine distribution promotes the evolution of vaccine escape"

### Supplementary Information

Philip J Gerrish[1,2,3,*], Fernando Saldaña[4], Benjamin Galeota-Sprung[5], Alexandre Colato[6], Erika E Rodriguez[7], Jorge X Velasco Hernández[4]

1. Department of Biology, University of New Mexico, Albuquerque NM 87131 USA
2. Theoretical Biology & Biophysics, Los Alamos National Lab, Los Alamos NM 87545 USA
3. Instituto de Ciencias Biomédicas, Universidad Autónoma de Ciudad Juárez, México
4. Instituto de Matemáticas, Universidad Nacional Autónoma de México, Juriquilla, México
5. Department of Biology, University of Pennsylvania, Philadelphia PA 19104 USA
6. Departamento Ciências da Natureza, Matemática e Educação, Universidade Federal de São Carlos, Araras SP, Brazil
7. Instituto de Ciencias Básicas e Ingenieria (ICBI), Universidad Autónoma del Estado de Hidalgo, Pachuca, México

* To whom correspondence should be addressed:

Philip Gerrish
167 Castetter Hall, MSC03 2020
1 University of New Mexico
Albuquerque, NM 87131-0001

### Model of vaccine escape emergence

For the convenience of the reader, we re-write parts of the basic model here so as to have everything in one place.

#### Fundamental assumption and its justification

Our approach is based on an assumption that an escape mutant will always be advantageous in a vaccinated population (SM). This assumption allows us to focus exclusively on the timing of the first infection event in which a new host is infected with an escape mutant, which we call an "escape-infection" event.

As a first approximation to justifying this assumption, we imagine a population with $S$ susceptible hosts (unvaccinated) and $V$ vaccinated hosts. Under a simple SIR model, assuming that unmutated "wildtype" virus can only infect unvaccinated hosts, the basic reproductive number of the wildtype virus is:

$$R_0 = \frac{\beta S}{\gamma}$$

where $\beta$ and $\gamma$ are infection and recovery rates, respectively. A vaccine escape mutant, however, in the same population has basic reproductive number:

$$R_0' = \frac{\beta(S + \sigma V)}{\gamma} = R_0 + \frac{\beta \sigma V}{\gamma}$$

where $\sigma$ is the probability that an escape mutant successfully infects a vaccinated host; this quantity might be called "escape efficacy" and is the complement of "vaccine efficacy" (see the next section). From which we conclude that the escape mutant has an advantage over wildtype ($R_0' > R_0$).

#### The model

Our model is described by the following equations:

$$\begin{aligned} \dot{S}_j &= -\sum_k^n \beta_{kj} I_k S_j - \phi_j S_j, \qquad \dot{I}_j = \sum_k^n \beta_{jk} I_j S_k - (\gamma + U) I_j, \\ \dot{V}_j &= \phi_j S_j, \qquad \dot{E}_j = U I_j - \gamma E_j, \qquad \dot{R}_j = \gamma (I_j + E_j), \end{aligned} \tag{1}$$

where $S_j$, $V_j$, $I_j$, $E_j$, and $R_j$ are the fraction of the population that are susceptible, vaccinated, infected, infected with escape mutant, and recovered, respectively, in Patch $j$; $\beta_{ij}$ is the transmission rate from Patch

$i$ to Patch $j$ ($\beta_{jj}$ is the transmission rate within Patch $j$); $\phi_j$ is vaccination rate in Patch $j$; $\gamma$ is recovery rate; $U$ is a composite per-host mutation rate from wildtype virus to escape mutant virus; $n$ is number of patches; dots indicate time derivatives.

We note the absence of a contagion term in the equation for $E_j$. This term is not needed for our purposes because our focus is only on the first escape-infection event – a discrete event. Furthermore, this term can lead to erroneous results because ours is a continuous model: a contagion term would allow for transmission to fractions of individual hosts that can erroneously amplify the vaccine escape mutant prior to the first escape-infection event.

Here, we assume there are only two patches, $n = 2$ and $j \in [1, 2]$, and that vaccination only happens in Patch 1, $\phi_2 = 0$. Our more complex models and detailed simulations are described in the next sections.

We define random variable $T_{ij}$ as the time of the first infection event in which a new host in Patch $j$ is infected by an escape mutant that arose in Patch $i$. Such infection events occur with rate $r_{ij}(t) = \beta_{ij}E_i(t)(S_j(t)+\sigma V_j(t))$, where $\sigma$ allows for varying levels of escape reflecting the observed spectrum of partial immunity against different variants ranging from no escape $\sigma = 0$ to full escape $\sigma = 1$. Put differently, $\sigma$ is the probability that an escape mutant successfully infects a vaccinated host; this quantity might be called "escape efficacy" and is the complement of "vaccine efficacy".

We will assume intra-patch transmission rates are equal, $\beta_{jj} = \beta$, and inter-patch transmission rates are equal, $\beta_{ij}|_{i\neq j} = \beta_\times$. We let $\beta_\times = \lambda\beta$ and we assume $\lambda \ll 1$ to reflect the fact that inter-patch transmission will typically be much less frequent than intra-patch transmission. We define random variable $T_f$ as the time at which the last infected individual recovers.

### Vaccine escape risk indicators

The three quantities of interest are:

1. $p = \mathbb{P}\{T_{11} > T_{21} \mid T_{21} < T_f \vee T_{11} < T_f\}$, the probability that vaccine escape in Patch 1 comes not from Patch 1 but from neighboring unvaccinated Patch 2, conditioned on vaccine escape emerging in Patch 1 from one of the two patches,

2. $f = \mathbb{P}\{T_{21} < T_f \vee T_{11} < T_f\}/\mathbb{P}\{T_{11} < T_f\}$, the factor by which the probability of vaccine escape in Patch 1 is increased by having neighboring unvaccinated Patch 2.

3. $\varepsilon = \mathbb{P}\{T_{11} < T_f \vee T_{12} < T_f \vee T_{21} < T_f \vee T_{22} < T_f\}$, the total probability of vaccine escape in the two patches as a function of vaccine distribution between the two patches.

These quantities are functions of the $r_{ij}(t)$ as defined above and in the main text. Escape-infection events form non-homogeneous Poisson processes with intensity functions $r_{ij}(t)$.

1. We first note that:

$$\mathbb{P}\{T_{11} > T_{21} \bigcap T_{12} < T_f \vee T_{11} < T_f\} = \int_0^\infty r_{21}(t) \exp\left(-\int_0^t r_{21}(u) + r_{11}(u)du\right) dt$$

Next, we note that:

$$\mathbb{P}\{T_{12} < T_f \vee T_{11} < T_f\} = 1 - \exp\left(-\int_0^\infty r_{21}(u) + r_{11}(u)du\right)$$

Giving:

$$\begin{aligned} p &= \mathbb{P}\{T_{11} > T_{21} \mid T_{12} < T_f \vee T_{11} < T_f\} = \frac{\mathbb{P}\{T_{11} > T_{21} \bigcap T_{12} < T_f \vee T_{11} < T_f\}}{\mathbb{P}\{T_{12} < T_f \vee T_{11} < T_f\}} \\ &= C \int_0^\infty r_{21}(t) \exp\left(-\int_0^t r_{21}(u) + r_{11}(u)du\right) dt \end{aligned} \tag{2}$$

where:

$$C = \left(1 - \exp\left(-\int_0^\infty r_{21}(u) + r_{11}(u)du\right)\right)^{-1}$$

2. The second quantity of interest is:

$$f = \frac{\mathbb{P}\{T_{12} < T_f \vee T_{11} < T_f\}}{\mathbb{P}\{T_{11} < T_f\}} = \frac{1 - \exp\left(-\int_0^\infty (r_{21}(t) + r_{11}(t))dt\right)}{1 - \exp\left(-\int_0^\infty r_{11}(t)dt\right)} \tag{3}$$

3. The third quantity of interest is:

$$\begin{aligned} \varepsilon &= \mathbb{P}\{T_{11} < T_f \vee T_{12} < T_f \vee T_{21} < T_f \vee T_{22} < T_f\} \\ &= 1 - \exp\left(-\int_0^\infty r_{11}(t) + r_{12}(t) + r_{21}(t) + r_{22}(t)dt\right) \end{aligned} \tag{4}$$

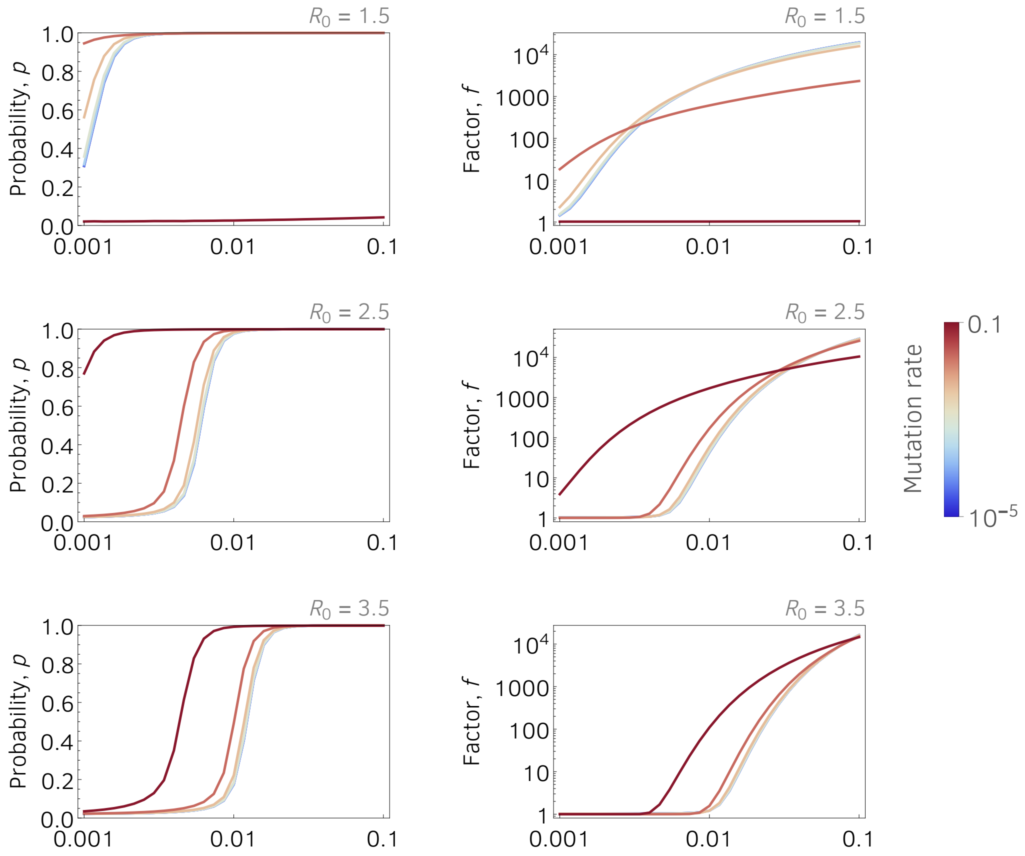


**Fig S1** | Parameters are: $\beta = \gamma R_0$, $\gamma = 0.1$, $\lambda \;=\; 0.02$, $N = 10^5$.

The third quantity of interest is the "vaccine escape factor", defined as the probability of vaccine escape relative to (i.e., divided by) the probability of vaccine escape when all the vaccine goes to one patch and none goes to the other (maximum vaccine disparity). This quantity is computed as follows:

$$\begin{aligned}\varepsilon \;&=\; k\mathbb{P}\{T_{11} < T_f \vee T_{12} < T_f \vee T_{21} < T_f \vee T_{22} < T_f\} \\ &=\; k\left(1 - \exp\left(-\int_0^\infty r_{11}(t) + r_{12}(t) + r_{21}(t) + r_{22}(t)dt\right)\right) \qquad (5)\end{aligned}$$

where $k$ is a constant insuring that $\varepsilon = 1$ under maximum vaccine disparity (when the vaccine all goes to one of the two patches).

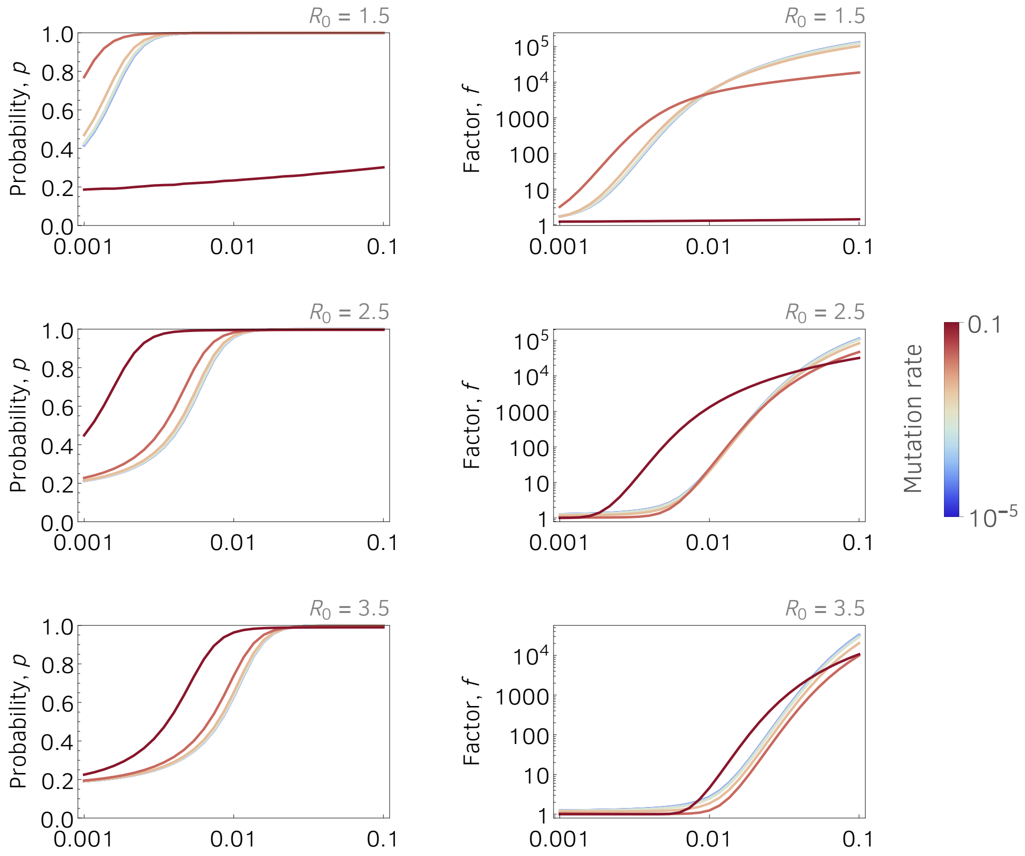


**Fig S2** | Parameters are: $\beta = \gamma R_0$, $\gamma = 0.1$, $\lambda = 0.2$, $N = 10^5$.

### Vaccine escape dynamics with natality/mortality and waning immunity

We present here a simple model to explicitly model escape mutant dynamics and find equilibrium points. Assume that the population is constant, normalized so that the independent variables always sum to one, with natality and mortality rate equal to $\mu$, and that the vaccine and the natural infection confer both temporal immunity for $1/\omega$ days on average. Let $S_i$, $V_i$, $I_i$, $E_i$ and $R_i$ denote the populations of susceptible, vaccinated, infected with the wildtype, infected with the escape mutant, and recovered (immune) individuals, respectively, in Patch $i$. Susceptible individual are vaccinated at a rate $\phi_i$ per unit time in Patch $i$. Susceptible individuals are infected by $I_i$ and $E_i$ individuals at rates $\beta S_i I_i$ and $\beta S_i E_i$, respectively, where $\beta$ is the effective contact/transmission rate. Vaccinated individuals can only be infected by the mutant vaccine escape strain at a rate $\sigma\beta V_i E_i$. Individuals infected by either of the two strains recover at the same rate $\gamma$. The model

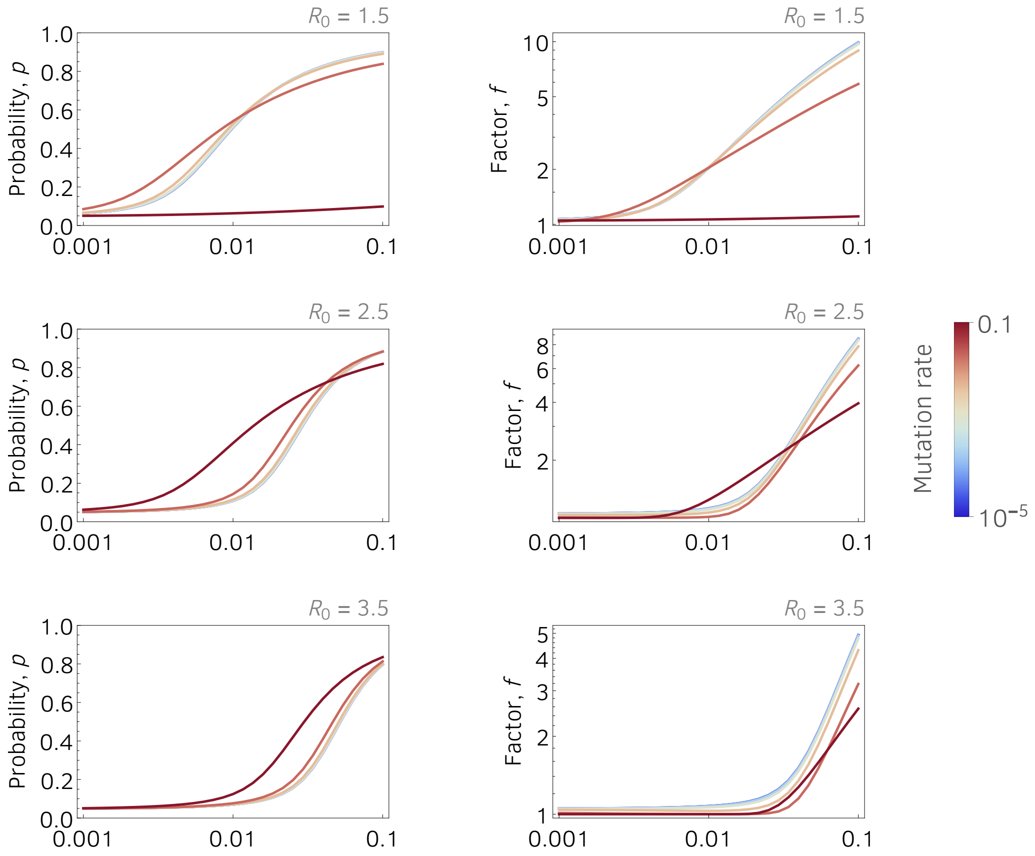


**Fig S3** | Parameters are: $\beta = \gamma R_0$, $\gamma = 0.1$, $\lambda\ =\ 0.05$, $N = 10^3$.

equations are then the following:

$$\begin{aligned}
S_i' &= \mu - S_i\beta_i(I_i + E_i) - (\mu + \phi_i)S_i + \omega(V_i + R_i),\\
V_i' &= \phi_i S_i - \sigma\beta V_i E_i - (\mu + \omega)V_i,\\
I_i' &= \beta S_i I_i - (\mu + \gamma)I_i,\\
E_i' &= \beta E_i(S_i + \sigma V_i) - (\mu + \gamma)E_i,\\
R_i' &= \gamma(W_i + E_i) - (\mu + \omega)R_i.
\end{aligned} \tag{6}$$

The initial conditions for this system are $S_i(0) = 1 - 1/N$, $V_i(0) = 0$, $I_i(0) = 1/N$, $E_i(0) = 0$ and $R_i(0) = 0$. $N$ is total population size.

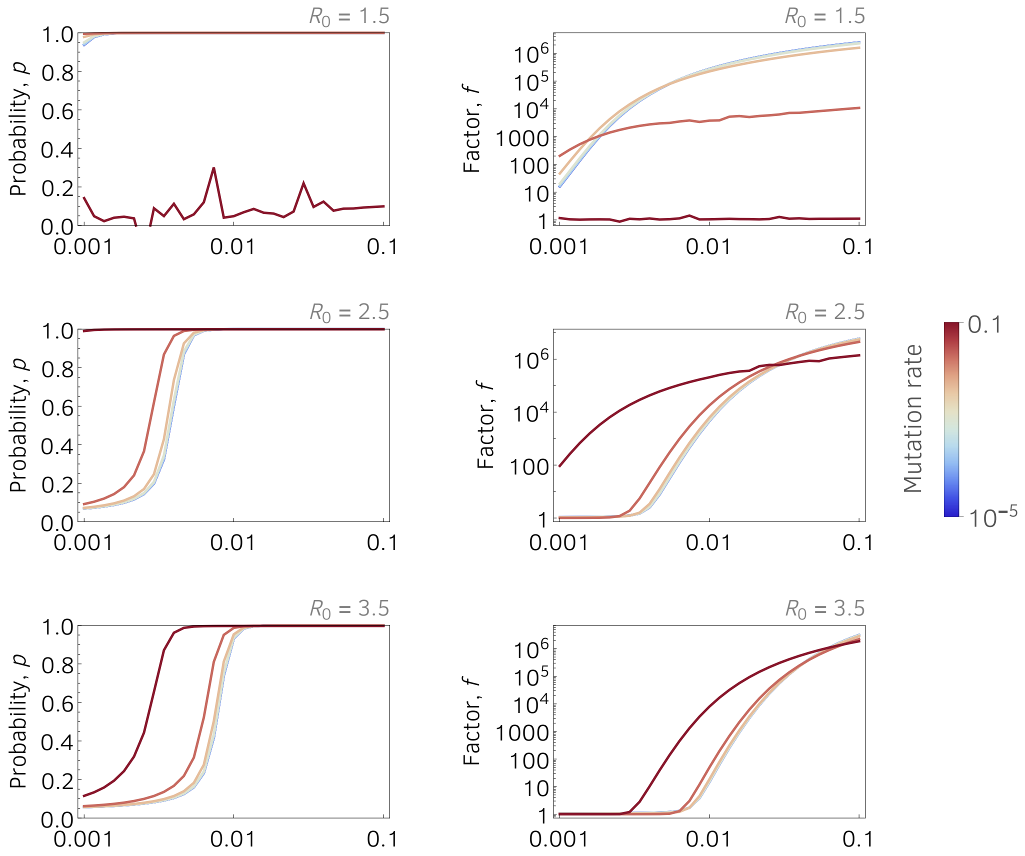


**Fig S4** | Parameters are: $\beta = \gamma R_0$, $\gamma = 0.1$, $\lambda = 0.05$, $N = 10^7$.

The disease free equilibrium is

$$S_* = \frac{\mu + \omega}{\mu + \omega + \phi}, \qquad V_* = \frac{\phi}{\mu + \omega + \phi},$$

with the other coordinates equal to zero. The basic reproductive number is the maximum of the following:

$$R_I = \frac{\beta(\mu + \omega)}{(\gamma + \mu)(\mu + \omega + \phi)}, \tag{7}$$

$$R_E = R_I + \frac{\sigma\beta}{\gamma + \mu}, \tag{8}$$

where the subindices $I$ and $E$ stand for the wildtype and escape mutant, respectively. Note that the wildtype reproductive number depends on the vaccine coverage $\phi$ and waning rate $\omega$ whereas the reproductive number for the mutant also depends on $\sigma$ the "escape efficacy" parameter.

*Support for our fundamental assumption:* Equation (8) clearly indicates that, at disease-free equilibrium, it

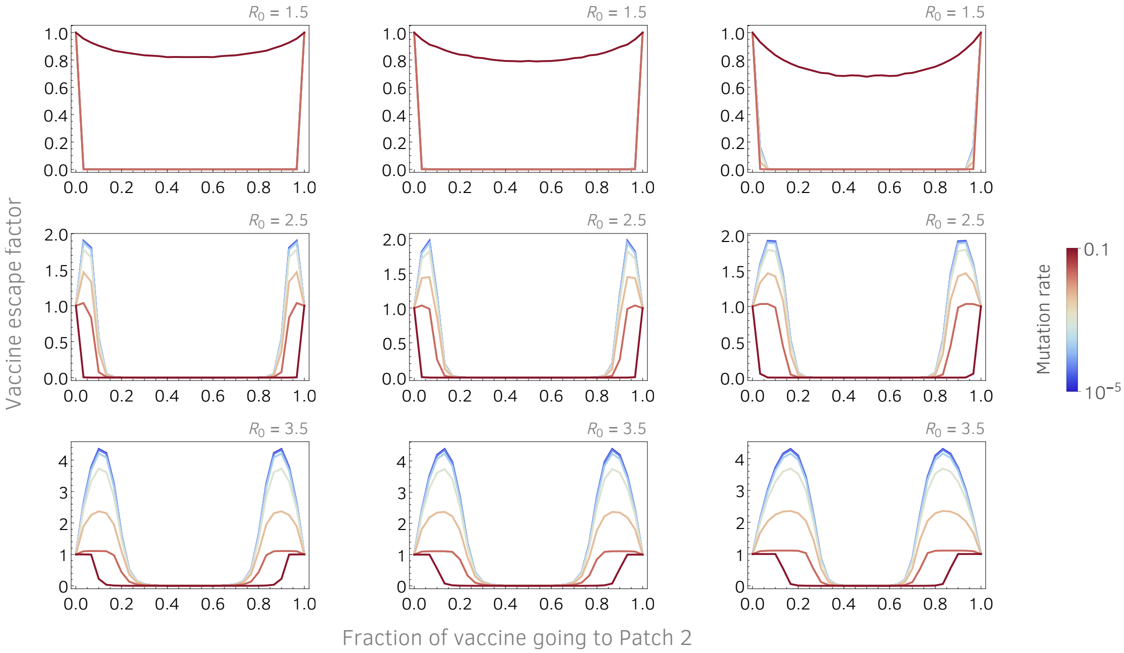


**Fig S5** | Vaccine escape factor, Eq (5), as a function the fraction of vaccine that goes to Patch 2. Vaccine escape factor is defined as vaccine escape probability divided by the escape probability when all vaccine goes to one of the two patches (maximum disparity). Parameters are: $\gamma = 0.1$, $\lambda = 0.02$, $N = 10^5$. *Left-hand column*: $V(0) = 0$, $\phi_1 + \phi_2 = 0.05$. *Middle column*: $V(0) = 0.2$, $\phi_1 + \phi_2 = 0.02$. *Right-hand column*: $V(0) = 0.6$, $\phi_1 + \phi_2 = 0.01$.

is always true that $R_E \geq R_I$, which again supports our underlying assumption that the escape mutation will be advantageous under vaccination.

### Incorporating escape mutation and migration as an ongoing discrete processes

We now introduce a modification of the above model: we introduce a discrete escape mutation process from wildtype to escape mutant, and a discrete migration process between patches. Here, at each time step, an escape mutation may appear in Patch $i$ with probability $UI_i$. An escape mutation, for example, that arises in Patch 2 will infect a susceptible host in Patch 1 with probability $\beta_{21}E_2(t)(S_1(t) + \sigma V_1(t))$.

In the numerical simulation of this system of equations, we consider two patches ($i \in \{1, 2\}$), with discrete inter-patch infection occurring at per-host rate $\beta_\times$, and discrete mutation from wildtype to escape mutant occurring at per-host rate $U$. We assume that there is vaccination in Patch 1 and it occurs at rate $\phi_1$, and that there is no vaccination in Patch 2, i.e., $\phi_2 = 0$.

In Fig S11, we see that the vaccinated Patch 1 was infected by an escape mutant coming from Patch 2,

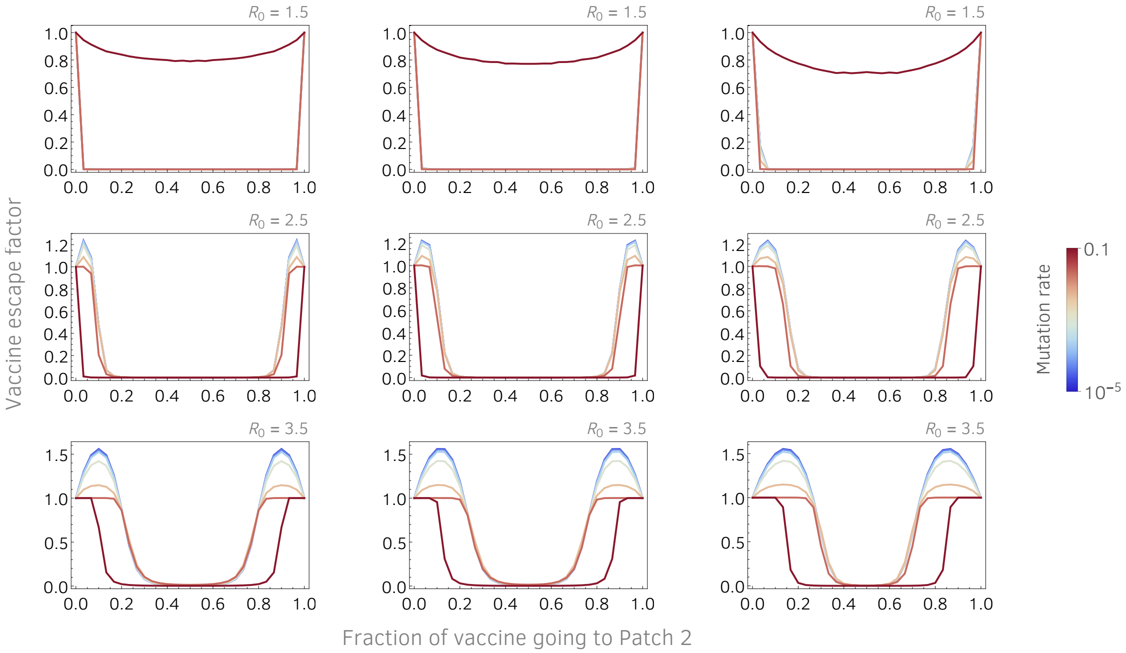


**Fig S6** | Vaccine escape factor, Eq (5), as a function the fraction of vaccine that goes to Patch 2. Vaccine escape factor is defined as vaccine escape probability divided by the escape probability when all vaccine goes to one of the two patches (maximum disparity). Parameters are: $\gamma = 0.1$, $\lambda = 0.2$, $N = 10^5$. *Left-hand column*: $V(0) = 0$, $\phi_1 + \phi_2 = 0.05$. *Middle column*: $V(0) = 0.2$, $\phi_1 + \phi_2 = 0.02$. *Right-hand column*: $V(0) = 0.6$, $\phi_1 + \phi_2 = 0.01$.

which subsequently reignited a new epidemic in Patch 1.

Figs S12 and S13 show the dynamics of the same representative simulation. Of special note is the observation that the first escape mutant to appear in Patch 1 came from Patch 2.

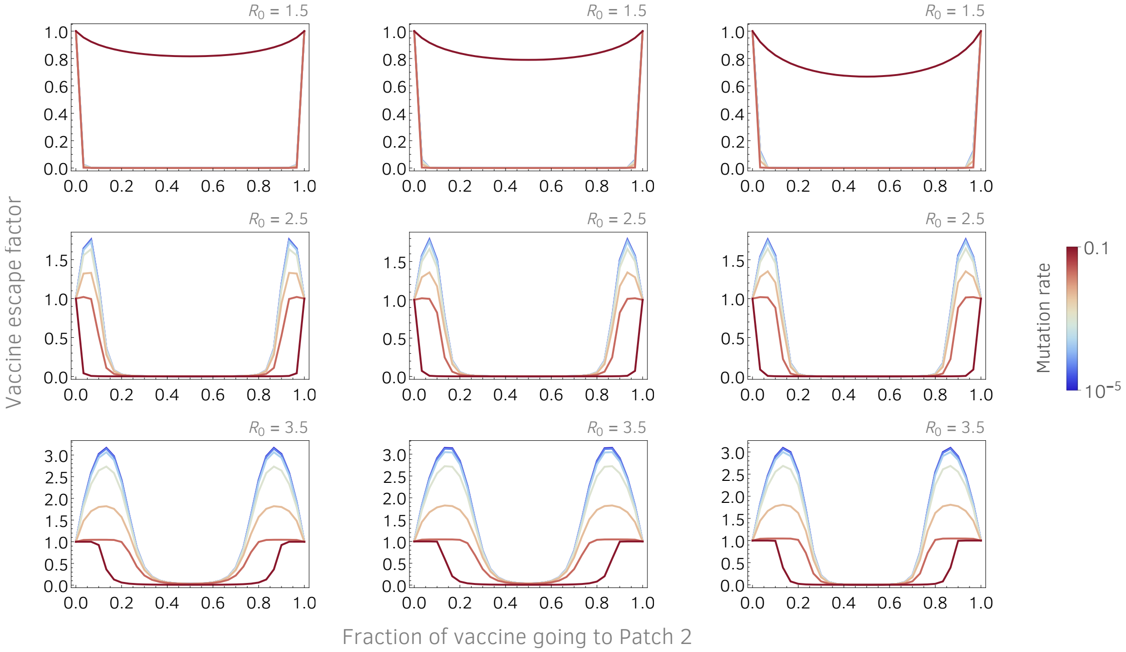


**Fig S7** | Vaccine escape factor, Eq (5), as a function the fraction of vaccine that goes to Patch 2. Vaccine escape factor is defined as vaccine escape probability divided by the escape probability when all vaccine goes to one of the two patches (maximum disparity). Parameters are: $\gamma = 0.1$, $\lambda = 0.05$, $N = 10^5$. *Left-hand column*: $V(0) = 0$, $\phi_1 + \phi_2 = 0.05$. *Middle column*: $V(0) = 0.2$, $\phi_1 + \phi_2 = 0.02$. *Right-hand column*: $V(0) = 0.6$, $\phi_1 + \phi_2 = 0.01$.

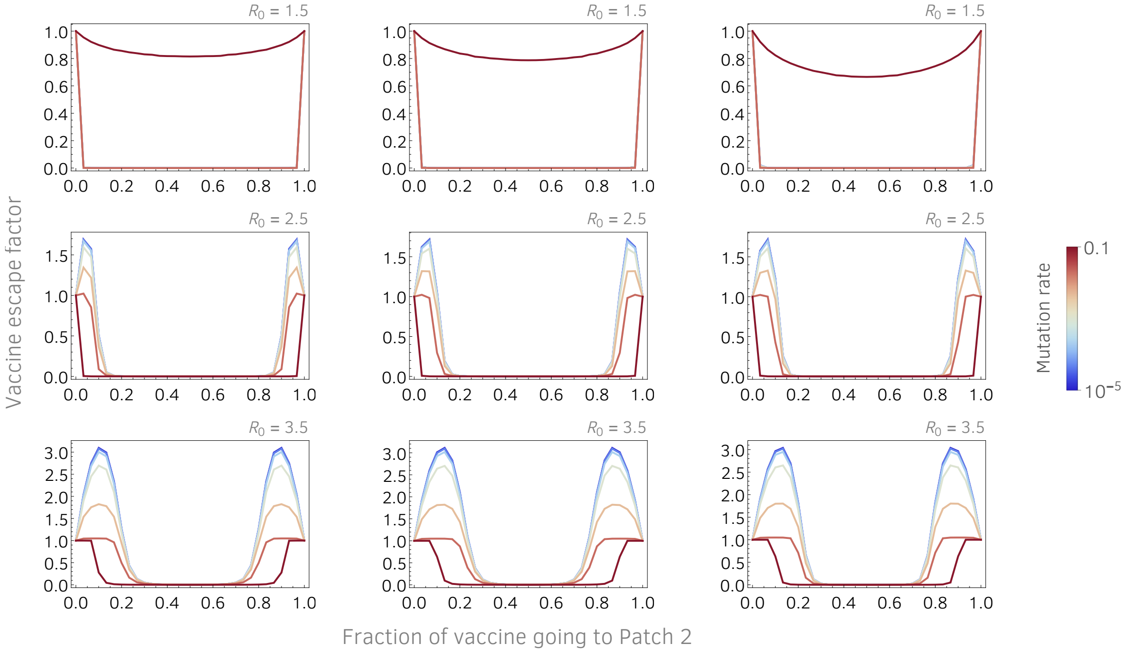


**Fig S8** | Vaccine escape factor, Eq (5), as a function the fraction of vaccine that goes to Patch 2. Vaccine escape factor is defined as vaccine escape probability divided by the escape probability when all vaccine goes to one of the two patches (maximum disparity). Parameters are: $\gamma = 0.1$, $\lambda = 0.05$, $N = 10^7$. *Left-hand column*: $V(0) = 0$, $\phi_1 + \phi_2 = 0.05$. *Middle column*: $V(0) = 0.2$, $\phi_1 + \phi_2 = 0.02$. *Right-hand column*: $V(0) = 0.6$, $\phi_1 + \phi_2 = 0.01$.

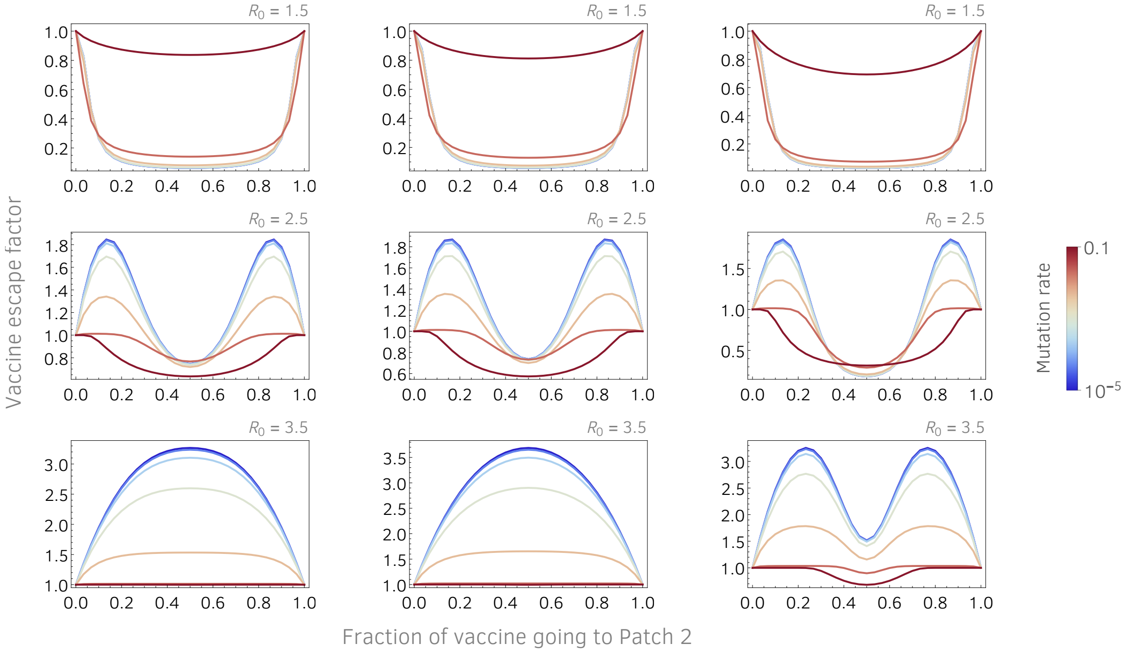


**Fig S9** | Vaccine escape factor, Eq (5), as a function the fraction of vaccine that goes to Patch 2. Vaccine escape factor is defined as vaccine escape probability divided by the escape probability when all vaccine goes to one of the two patches (maximum disparity). Parameters are: $\gamma = 0.1$, $\lambda = 0.05$, $N = 10^3$. *Left-hand column*: $V(0) = 0$, $\phi_1 + \phi_2 = 0.05$. *Middle column*: $V(0) = 0.2$, $\phi_1 + \phi_2 = 0.02$. *Right-hand column*: $V(0) = 0.6$, $\phi_1 + \phi_2 = 0.01$.

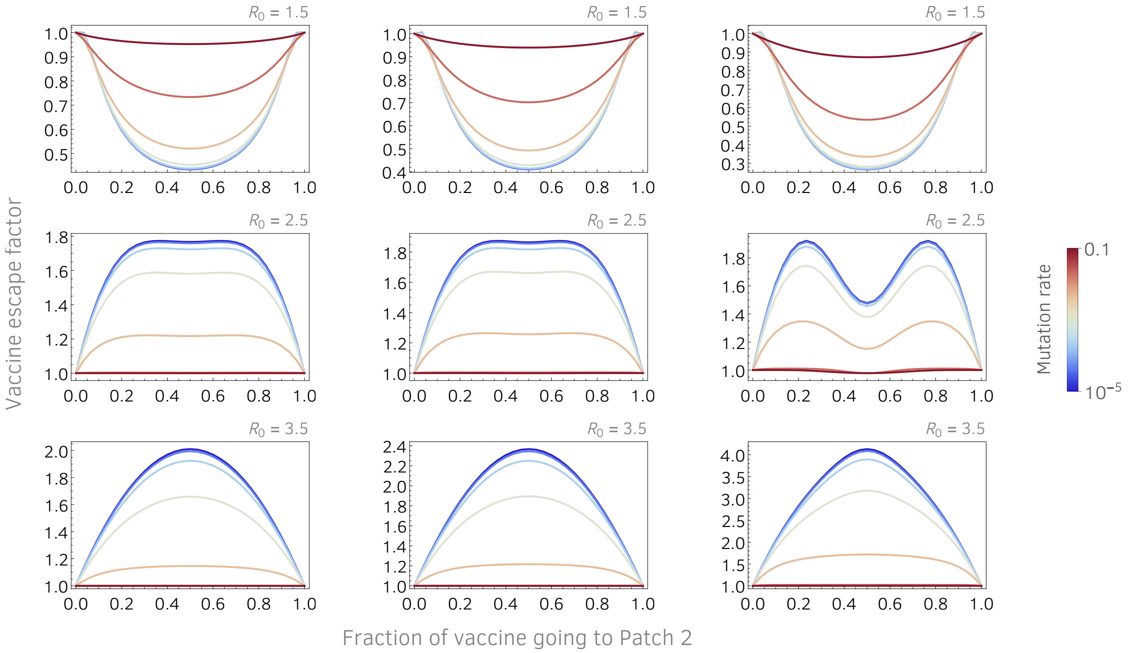


**Fig S10** | Vaccine escape factor, Eq (5), as a function the fraction of vaccine that goes to Patch 2. Vaccine escape factor is defined as vaccine escape probability divided by the escape probability when all vaccine goes to one of the two patches (maximum disparity). Parameters are: $\gamma = 0.1$, $\lambda = 0.05$, $N = 10^2$. *Left-hand column*: $V(0) = 0$, $\phi_1 + \phi_2 = 0.05$. *Middle column*: $V(0) = 0.2$, $\phi_1 + \phi_2 = 0.02$. *Right-hand column*: $V(0) = 0.6$, $\phi_1 + \phi_2 = 0.01$.

```
Out[•]= {{v: 2→1, 34.5}, {s: 1→2, 61.4}, {s: 1→2, 62.3},
  {s: 1→2, 76.8}, {s: 1→2, 77.9}, {s: 1→2, 80.9}, {s: 1→2, 93.9}}
```

**Fig S11** | Table of migration events of escape mutations in one representative simulation. "v" indicates that the migration event resulted in the infection of a vaccinated individual by an escape mutant. "s" indicates that the migration event resulted in the infection of a susceptible individual by an escape mutant. $1 \to 2$ indicates that the escape mutation originated in Patch 1 and migrated to Patch 2. $2 \to 1$ indicates that the escape mutation originated in Patch 2 and migrated to Patch 1. The number that follows indicates the time at which the migration occurred.

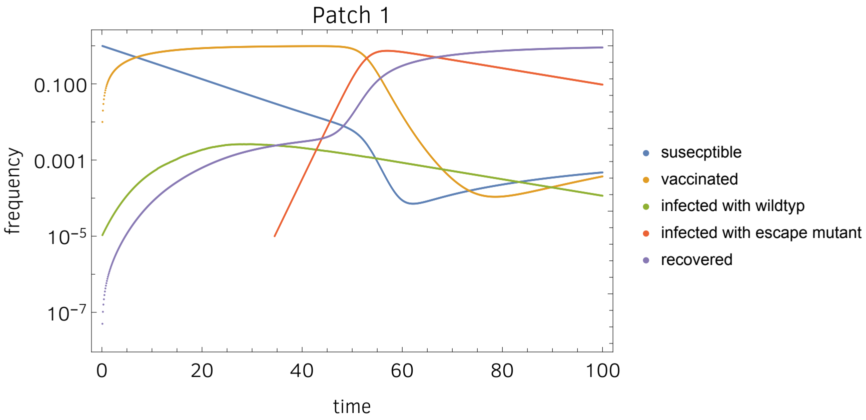


**Fig S12** | Dynamics of epidemic in Patch 1. We plot a representative simulation. Escape mutant arose in Patch 2 and migrated to Patch 1 at generation 34.5, as indicated in Fig S11

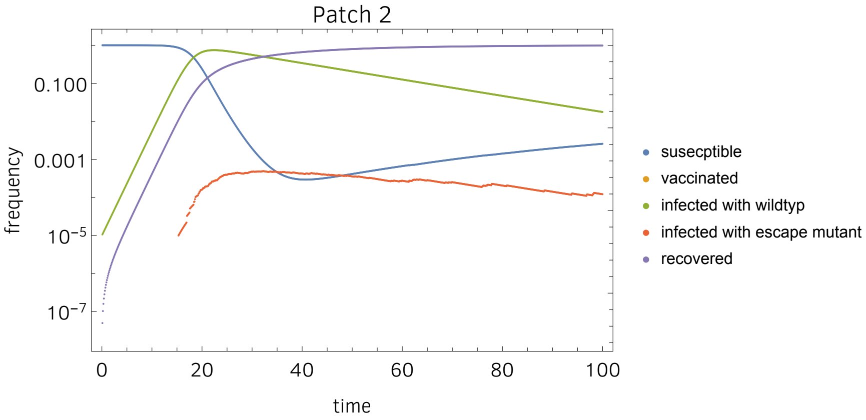


**Fig S13** | Dynamics of epidemic in Patch 2. We plot a representative simulation. Escape mutant arose in Patch 2 and migrated to Patch 1 at generation 34.5, as indicated in Fig S11

**Distribution of time until first escape-infection event: full model with natality/mortality and waning immunity**

An adaptation of the above model, Eq (6), to determine the distributions of times until the first escape-infection event is given by the following system of equations:

$$\begin{aligned}
S_i' &= \mu - \sum_k^n \beta_{ki} I_k S_i - (\mu + \phi_i) S_i + \omega(V_i + R_i),\\
V_i' &= \phi_i S_i - (\mu + \omega) V_i,\\
I_i' &= \sum_k^n \beta_{ik} I_i S_k - (\mu + \gamma) I_i - U I_i,\\
E_i' &= U I_i - (\mu + \gamma) E_i,\\
R_i' &= \gamma(I_i + E_i) - (\mu + \omega) R_i.
\end{aligned} \tag{9}$$

The effects of the new factors introduced here, natality/mortality and waning immunity, are evaluated in Figs S14 and S15. We note: since we are already modeling the the evolution of the virus, it might reduce the potential for confusion to interpret waning immunity here as a property of the host and not as a consequence of the evolution of the virus.

### Small populations can go the other way

We call attention to Fig S9 and especially Fig S10, which show that in small populations and high $R_0$, vaccine disparity between the two patches can either have no effect at high mutation rates or a favorable effect at low mutation rates. This observation may perhaps be intuited by considering that low mutation rates and small population sizes will make mutants scarce and selection weak. Vaccine disparity between these small populations may thus be optimal for suppressing vaccine escape mutants because the won't appear in the vaccinated compartment due to lack of wildtype replication, and they may appear in the unvaccinated population but at a rate not much different from both compartments under equal vaccination. Vaccine disparity in small populations at low mutation rates thus has the effect of simply eliminating one of the compartments.

The implication is that highly granular vaccine disparities – large adjacent populations such as cities, states or countries that differ in vaccine accessibility – most effectively promote the evolution of vaccine escape. As the disparities become less granular (finer-grained disparities), the effect appears to disappear, at least

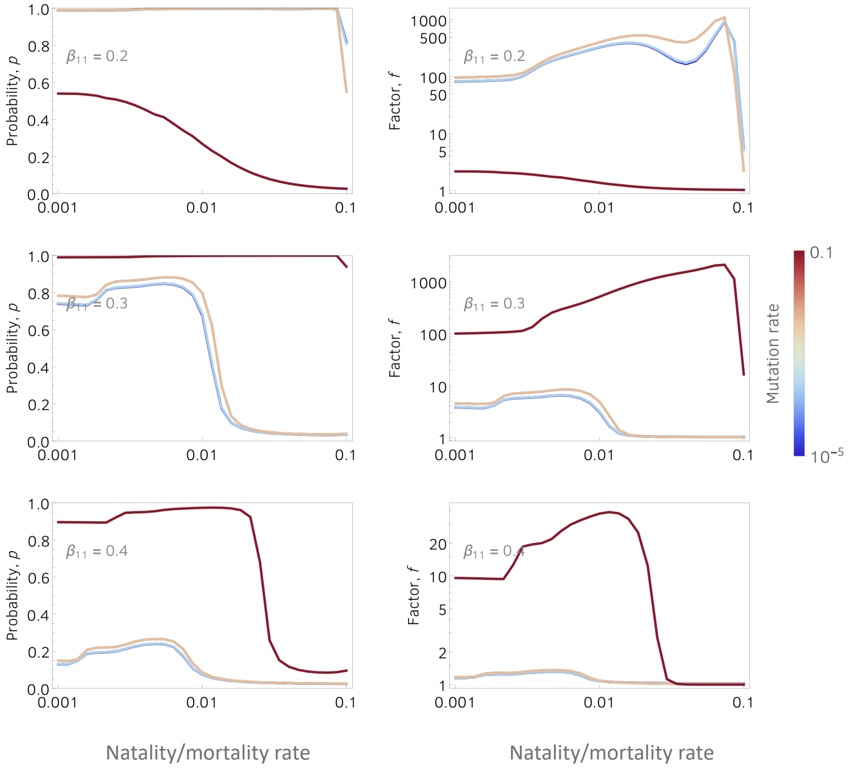


**Fig S14** | Parameters are: $\gamma = 0.1$, $\lambda = 0.1$, $\phi_1 = 0.02$, $\phi_2 = 0$.

for moderate to high $R_0$.

### Model parameters

Here we assess the ranges of the model parameters we use in our figures, and their appropriateness for modeling SARS-CoV-2. First, for the simple SIR model which is a special case of the vaccination model (1), the basic reproduction number is:

$$R_0 = (\beta/\gamma)N = \beta/\gamma \tag{10}$$

because $N = 1$. If we assume the average recovery time from covid-19 is 10 days, we have $\gamma = 1/10$ days is a realistic value for the recovery rate $\gamma$. Using these values, we can obtain $\beta$ to reflect a specific $R_0$. Estimations of $R_0$ for covid are between $2 - 4$, so fixing $R_0 = 2$ we obtain $\beta = 0.2$ is a realistic value and $\beta \in [0.1 - 0.4]$ is a realistic range assuming fixed $\gamma = 1/10$.

On the other hand, to obtain and approximation for a constant vaccination rate $\phi_1$, let us consider a normalized total population $N(t)$ where at no individuals has been vaccinated at the initial time. Assuming

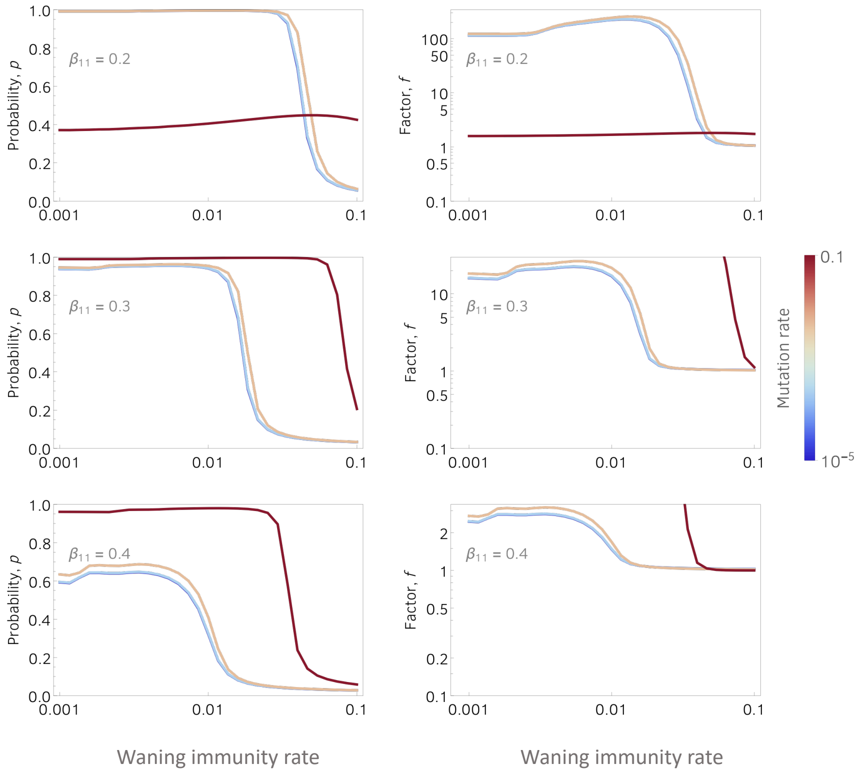


**Fig S15** | Parameters are: $\gamma = 0.1$, $\lambda = 0.1$, $\phi_1 = 0.02$, $\phi_2 = 0$.

that the vaccination rate $\phi_1$ is proportional to population size, we have $N'(t) = -\phi_1 N(t)$, $N(0) = 1$. A direct computation allow us to obtain $N(t) = \exp(-\phi_1 t)$, hence, the fraction of vaccinated individuals at time $t$, that is, the immunization coverage, $C$, is $C(t) = 1 - \exp(-\phi_1 t)$. For a fixed time horizon $T$, we have

$$C(T) = 1 - \exp(-\phi_1 T) \text{ or } \phi_1 = -\ln(1 - C(T))/T. \tag{11}$$

Considering a very optimistic case in which health authorities achieve a vaccination coverage $C(T) = 90\%$ of the population in $T = 100$ days, we obtain that the constant vaccination rate $\phi_1 = 0.023$. We explore vaccination rates on a logarithmic scale from $\phi_1 = 0.001$ than $\phi_1 = 0.1$.

### Description of agent-based Evolutionary Epidemiology simulations

We simulate a population of hosts, each of which can be infected by a population of virus, which is also simulated. The host population is subdivided into compartments, only some of which receive vaccination at per-capita vaccination rate $\phi_1$.

#### Time

- The unit of time throughout is the *viral generation.*

#### Virus properties

- Viral fitness. Viral fitness $w$ is a composite parameter with two components, defined as:

$$w = w_n + w_a$$

  where $w_n$ is the *non-antigenic fitness* or "intrinsic" fitness, and $w_a$ is the *antigenic fitness.*

- Non-antigenic fitness. This is the intrinsic competitive fitness of the virus that is not related to its antigenic properties. In the simulations, this fitness is represented by a continuous variable, whose value is passed from parent to offspring – sometimes changed in the process due to mutation (see below). Initially, all virions have non-antigenic fitness equal to zero.

- Antigenic fitness. This component of viral fitness is actually dependent on the antigenic configuration of the virus ($V$) *in relation to* the immune configuration of the host ($H$). [Employing this notation which will be clarified later, we can write $w_a = w_a(V, H)$.] Specifically, antigenic fitness is a function of the within-host Hamming distance between a virion's epitope(s) and host antibodies. The function we chose to map this Hamming distance, $d = d(V, H)$, onto $w_a$ is a logistic function:

$$w_a = \frac{w_{max}}{1 - e^{-rd} + e^{r(d_{1/2} - d)}}$$

  where $w_{max}$ is the maximum fitness, $r$ determines the "curviness" of the function (larger $r$ means a sharper rise to $w_{max}$), and $d_{1/2}$ is the distance at which the function is exactly $\frac{1}{2}w_{max}$. This function is plotted in Fig S16, the parameters for which were used in one set of simulations.

- Antigenic configuration. This is the part of the virus that is immuno-reactive and is a major component of a virus's overall fitness. In the simulations, it is represented by a "bit-string", that is, a string of

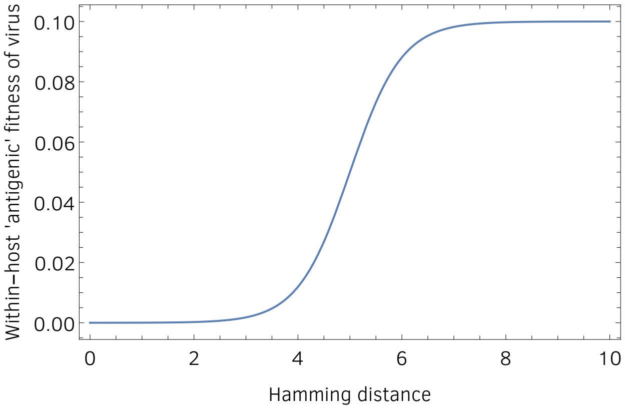


**Fig S16 | Antigenic component of viral fitness** as a function of the Hamming distance between the antigenic configuration of a virion and the immune configuration of its host. Because virions can have different antigenic configurations and because these configurations can undergo mutation, the viral antigenic configuration can evolve within-host to be increasingly distinct from the host's immune configuration.

ones and zeros. A bit-strings is compactly encapsulated by one or more 64-bit integers whose binary representation is the virion's antigenic bit-string. Each 64-bit integer models one viral immuno-reactive epitope; virions may have one or more epitopes. Each such epitope gets passed from parent to offspring virion, with possible mutation happening in the process (see below). The first virion infecting the first host has a bit-string of all zeros.

- Within-host viral population size. Here we make two, perhaps unrealistic, assumptions: 1) we assume that the population size is constant from the moment of infection and remains constant for the duration of the infection which is set at 100 viral generations. 2) we assume that the within-host population size is on the order of hundreds or perhaps thousands of virions; this is of course orders of magnitude smaller that real within-host viral populations. While each of these two assumptions, by itself, is unrealistic, an argument can be made that the two of these assumptions *taken together* can constitute a somewhat realistic *pair* of assumptions.

  To make this argument, 1) we appeal to the population-geneticist's "trick" of assuming that the population in question remains at a constant size that is stochastically equivalent to the real population which varies in size, called the *effective population size*; 2) we employ the fact that a very large population that repeatedly undergoes very small bottlenecks, as viral populations do (growing to large numbers within hosts but being subject to very small bottlenecks upon transmission to a new host), has an effective population size that is much closer to the bottleneck size than to the large inter-bottleneck size.

#### Within-host evolution of virus

- Replication and selection. Replication and selection is modeled as a biased Wright-Fisher process with mutation, which works as follows. Every viral generation, a new population of viral offspring is created simply as a Multinomial sample of the parent population. The multinomial sampling probability for parent $i$ is simply $w_i/\bar{w}$, where $w_i$ is the fitness of parent $i$ and $\bar{w}$ is mean fitness of the parent population. In this step, fitness and antigenic configuration are passed from parent to offspring virion with 100% fidelity; mutation occurs after each such replication step, as follows.

- Mutation.

  Mutation follows and is independent of each replication step.

  1. *Non-antigenic fitness.* After the viral population has replicated as described above, each offspring virion receives a Poisson-distributed number of new deleterious mutations with mean $U$ and a Poisson-distributed number of beneficial mutations with mean $\mu$. It is generally the case that $U \gg \mu$. The fitness of an offspring virion is calculated as:

  $$w_o \;=\; w_p \;+\; \sum_{i=1}^{B} S_b \;-\; \sum_{i=1}^{D} S_d$$

     where $w_o$ and $w_p$ are the fitnesses of the offspring and parent virions, respectively. The number of new beneficial mutations, $B$, is a Poisson random variable with mean $\mu$. The number of new deleterious mutations, $D$, is a Poisson random variable with mean $U$. The fitness effect of each new beneficial mutation, $S_b$, is an exponential random variable with mean $s_b$. The fitness effect of each new deleterious mutation, $S_d$, is an exponential random variable with mean $s_d$.

  2. *Antigenic configuration.* The antigenic configuration is also passed from parent to offspring virion. After the viral population has replicated as described above, each offspring virion may experience mutation in its antigenic configuration, as follows. The antigenic configuration of the $i^{th}$ epitope of an offspring virion is computed as follows:

  $$V_o(i) \;=\; V_p(i) \;\oplus\; \sum_{i=0}^{63} \Lambda 2^i$$

     where $V_o(i)$ and $V_p(i)$ are integers representing the antigenic configurations of the $i^{th}$ epitope of offspring and parent virions, respectively, and $\oplus$ is the bitwise exclusive OR operator. $\Lambda$ is a Bernoulli random variable with mean $\lambda$, the "per-nucleotide" epitope mutation rate.

**Host properties**

- Immune configuration. Each host is randomly assigned an immune configuration at the outset. This is achieved by assigning each host a bit-string for each epitope-specific antibody as follows:

$$H(i) = \sum_{i=0}^{63} \Delta 2^i$$

where $H(i)$ is an integer representing the immune configuration of the antibody specific to the $i^{th}$ viral epitope, of a given host. $\Delta$ is a Bernoulli random variable with mean $\delta$, a measure of the initial immunological diversity of the host population. A host's immune configuration can be modified: a) by infection with the virus, or b) by vaccination.

- Pairwise contact rate. At each unit of time (each viral generation), a number of pairwise contacts between hosts is generated at random. For any given pair generated, if one host is infected and the other is not, then there exists the possibility of a transmission event (see *Transmission* below). This pairwise contact rate is calculated to insure that $R_0 > 1$.

- Subdivided population. The host population is divided into two or more subpopulations, some of which receive vaccination at rate $\phi_1$; the others receive no vaccination.

- Status (susceptible, infected, vaccinated, recovered). Initially, there is only one infected host. All others are susceptible. Our simulations are different than most ODE models, in that the status of a host is not "prescribed" by the labels susceptible, infected, vaccinated or recovered, but is instead determined entirely by the immune configuration of the host in relation to the antigenic configuration of the circulating virus. Hence, the lines separating the traditional labels are blurred, and the status of a host can change over time, depending on vaccination and the evolution of the virus.

**Transmission**

- At each unit of time (each viral generation), a number of pairwise contacts between hosts is generated at random.

- With a certain low probability, prescribed by the parameter "mixrate", the two hosts are from two different compartments. Otherwise, the two hosts are from the same compartment.

- For any given pair generated, if one host is infected and the other is not, then there exists the possibility of a transmission event. The probability of transmission happening is a function of the mean Hamming distance between the antigenic configurations of a small number of virions chosen at random from the infected host (the inoculum) and the immune configuration of the uninfected host: if that distance is small, this means that the host's immune system recognizes the virus, and the probability of transmission is low; if that distance is large, on the other hand, this means that the host's immune configuration is very different from the virus's antigenic configuration, implying that the host's immune system will not recognize the virus, and the probability of transmission is large.[1]

- Again, the function we chose to map this Hamming distance, $d = d(V, H)$, onto probability of infection, $p_{inf}$ is a logistic function:

$$p_{inf} \;=\; \frac{p_{max}}{1 - e^{-rd} + e^{r(d_{1/2}-d)}}$$

where $p_{max}$ is the maximum probability of infection, $r$ determines the "curviness" of the function (larger $r$ means a sharper rise to $p_{max}$), and $d_{1/2}$ is the distance at which the function is exactly $\frac{1}{2}p_{max}$. This function is plotted in Fig S17, the parameters for which were used in one set of simulations.

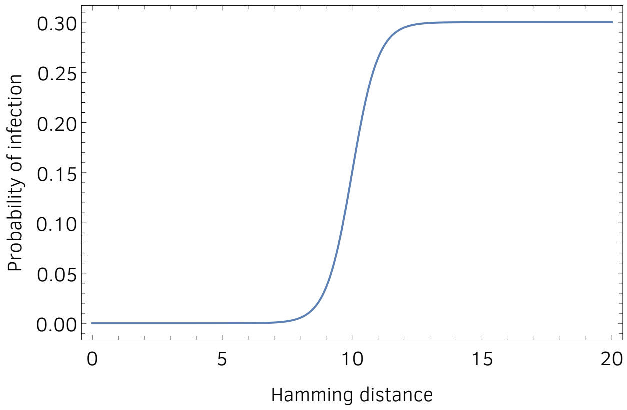


**Fig S17 | Probability of infection** as a function of the Hamming distance between the consensus viral epitope sequence in the inoculum and the immune antibody sequence.

[1]We are aware that a more realistic model would make protection against the virus an *increasing* function of Hamming distance between host and virus, because this would more realistically reflect complementary binding; operationally, however, ours is an equivalent approach.

**Infection**

- Upon successful transmission, a within-host viral population is established. This population is immediately of size $N_w$, the effective population size, and consists of a handful of viral genotypes that were present in the *inoculum.* The number of virions present in the inoculum is the "transmission bottleneck size" and is prescribed by the parameter $n_b$.

- Only at the end of the infection period, the immune configuration of the host is adjusted to closely match the antigenic configuration of the infecting virus. The within-host evolution of the virus is thus little affected by host immunity. This reflects the observation that transimissibility of SARS-CoV-2 peaks around the time of onset of symptoms [1] whereas a robust antibody response does not develop until roughly ten days after the onset of symptoms [1, 2]. The shift in host immune configuration at the end of the infection cycle is what will protect the host in the future from reinfection. If the circulating virus evolves rapidly, however, reinfection is possible. Depending on viral evolution rate, therefore, our simulations reflect an SIR model (low evolution rate) or an SIRS model (high evolution rate).

- The virus evolves within each host through standard mutation and selection processes.

**Vaccination**

- Vaccination occurs in designated *vaccinated compartments* at per-capita rate $\phi_1$.

- The vaccine closely matches the antigenic configuration of the initial virion that infected the first infected host. Namely, it closely matches a bit-string of all zeros.

- Upon vaccination, the immune configuration of the vaccinated host is adjusted to closely match the antigenic configuration of the vaccine. This close match is what protects the host from infection. If the circulating virus evolves rapidly, however, infection is possible. If infection of a vaccinated host happens, the viral variant infecting the host is designated a *vaccine escape mutant.*

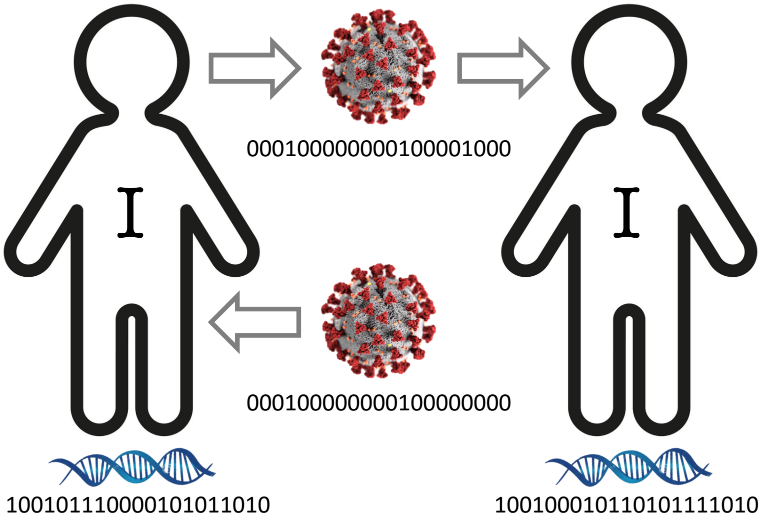


**Fig S18 | Viral transmission and evolution in a population of unvaccinated hosts**. The infecting virus (lower middle) has an epitope sequence very close to all zeros. It is able to infect the host on the left because the Hamming distance between the viral epitope (sequence below infecting virus) and host antibody (sequence on lower left) is large, so the host's immune system does not recognize the virus. The host on the left then transmits a slightly mutated virus (upper middle) to the host on the right. Again, this secondary infection is possible because the Hamming distance between the viral epitope (sequence below transmitted virus) and host antibody (sequence on lower right) is large, so the host's immune system does not recognize the virus.

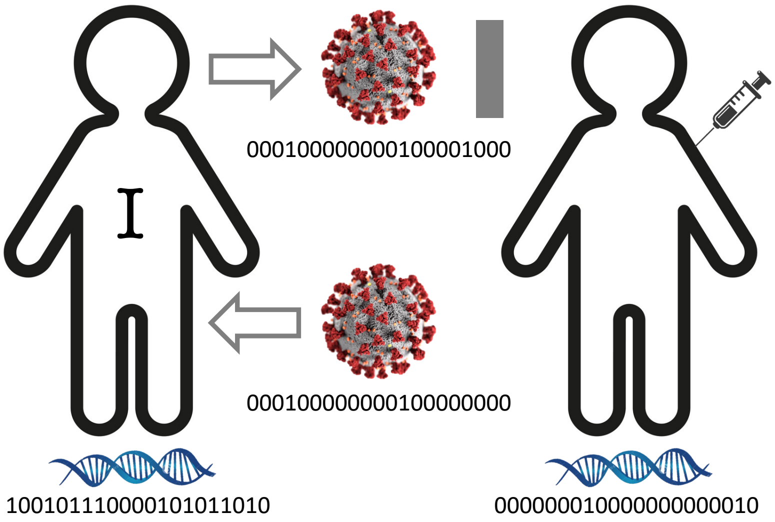


**Fig S19 | Viral transmission and evolution in a host population with vaccination**. Same as Fig S18, but here the host on the right was vaccinated before exposure to the virus. Vaccination is reflected by the fact that the antibody sequence of the vaccinated host (lower right) is very close to all zeros and very close to the epitope sequence of the virus to which he/she is exposed. Consequently, the vaccinated host's immune system recognizes the virus and the host is protected.

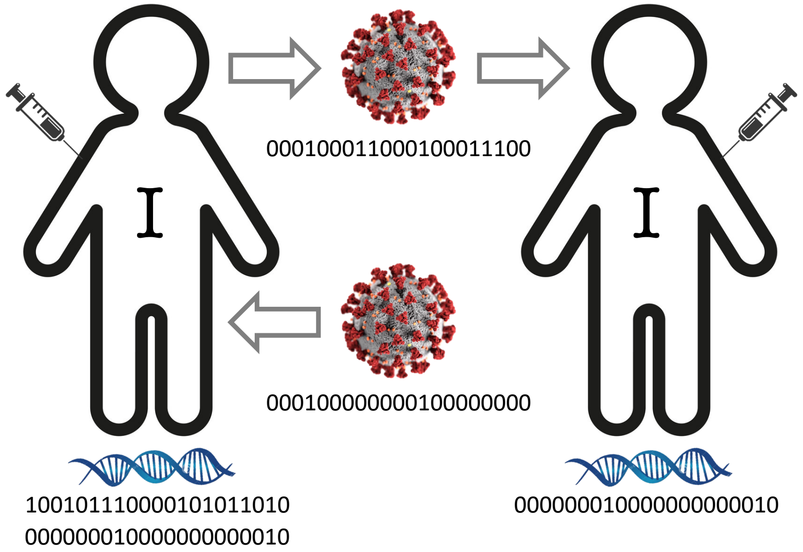


**Fig S20 | Viral transmission and evolution of vaccine escape in a population of vaccinated hosts**. Same as Fig S19, but here the host on the left is either: 1) vaccinated while infected, or 2) treated with immunologically similar monoclonal antibodies while infected. In either case, the virus will infect the unprotected host prior to vaccination or antibody therapy. The subsequent administration of vaccination or antibody therapy will then have the effect of creating *soft selection* for vaccine escape mutants in an already large within-host viral population. This is a recipe for the facile evolution of vaccine escape. When the escape mutant is then passed on to the next vaccinated host, it is likely to infect that host despite being vaccinated.

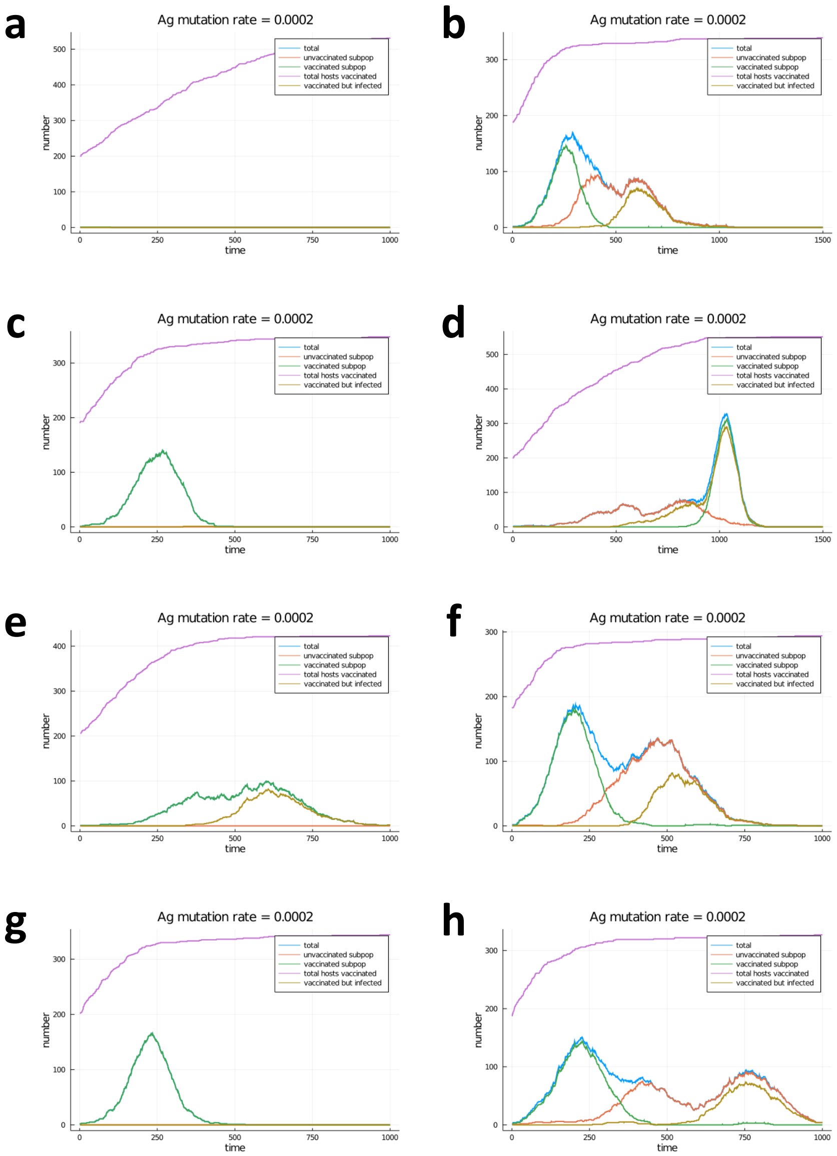


**Fig S21 | Comparing one- and two-compartment simulations**. Plots on the left derive from simulations with only one compartment with vaccination. Plots on the right derive from simulations with two compartments of equal size: one with vaccination and one with no vaccination. There was a low rate of migration between the two compartments. All simulations started out with a single infected host in each compartment and had exactly the same parameters. The green curve is number infected in the compartment with vaccination. The red curve is the number infected in the compartment with no vaccination. The blue curve is the total number infected. The purple curve is total number of hosts vaccinated (in the compartment with vaccination). Finally, the curve to pay special attention to is the gold-colored curve; this is the total number of hosts that are both vaccinated and infected; this is the indicator of how much vaccine escape there is. With the exception of plot **e**, vaccine escape does not appear in the simulations with a single vaccinated compartment (plots on left); however, significant vaccine escape appears in the simulations with one vaccinated compartment and one unvaccinated compartment (plots on right).
